# Preventing diabetic foot ulcers in low resource settings using Pedal Elevated Temperature Risk Assessment (PETRA)

**DOI:** 10.1101/2022.06.28.22276911

**Authors:** Kayla Huemer, Qingyue Wei, Srikar Nallan, Felix Jebasingh, Latha Palaniappan

## Abstract

Diabetic foot ulcers develop for up to 1 in 3 patients with diabetes. While ulcers are costly to manage and often necessitate an amputation, they are preventable if intervention is initiated early. However, with current standard of care, it is difficult to know which patients are at highest risk of developing an ulcer. Recently, thermal monitoring has been shown to catch the development of complications around 35 days in advance of onset. We seek to use thermal scans of patients’ with diabetes feet to automatically detect and classify a patient’s risk for foot ulcer development so that intervention may be initiated. We began by comparing performance of various architectures (backbone: DFTnet, ResNet50, and Swin Transformer) trained on visual spectrum images for monofilament task. We moved forward with the highest accuracy model which used ResNet50 as backbone (DFTNet acc. 68.18%, ResNet50 acc. 81.81%, Transformers: acc. 72.72%) to train on thermal images for the risk prediction task and achieved 96.4% acc. To increase interpretability of the model, we then trained this same architecture to predict two standard of care risk scores: high vs low-risk monofilament scores (81.8% accuracy) and high vs low-risk biothesiometer score (77.4% accuracy). We then sought to improve performance by facilitating the model’s learning. By annotating feet bounding boxes, we trained our own YoloV4 detector to automatically detect feet in our images (mAp accuracy of 99.7% and IoU of 86.%). By using these bounding box predictions as input to the model, this improved performance of our two classification tasks: MF 84.1%, BT 83.9%. We then sought to further improve the accuracy of these classification tasks with two further experiments implementing visual images of the feet: 1) training the models only on visual images (Risk: 97.6%, MF: 86.3%, BT: 80.6%), 2) concatenating visual images alongside the thermal images either early (E) or late (L) fusion in the architecture (Risk, E: 99.4%, L: 98.8% ; MF, E: 86.4%, L: 90.9%; BT, E: 83.9%, L: 83.9%). Our results demonstrate promise for thermal and visible spectrum images to be capable of providing insight to doctors such that they know which patients to intervene for in order to prevent ulceration and ultimately save the patient’s limb.

## 1 Introduction

Diabetic foot ulcers (DFU), or open sores on the bottom of the feet, develop for 1 in 3 patients with diabetes [1]. If left untreated, infection will soon threaten a patient’s life and may require amputation [2]. These amputations precede up to 80% of all lower-limb amputations [3], with one occurring every 20 seconds worldwide. Intervention at the earliest warning sign of a developing ulcer is key to preventing them from forming, but it’s difficult to know which patients are at highest risk of developing an ulcer [4]. In addition, barriers to access treatment in remote or low-resource extends the time until treatment, making it more likely patients will require an amputation. It is estimated that between of 50-90% of people living with diabetes are undiagnosed in rural areas of India [1].

The first indication of risk is the development of neuropathy. Standard neuropathy tests include the monofilament score (0-10 points, >6 is healthy) or biothesiometer score (0-50V, <25V is healthy). However, even with a positive neuropathy score, it may be months or years until an ulcer forms. In addition, current neuropathy test methods are time consuming, require training and AC power, are labor intensive, and outside the reach of low-resource healthcare settings.

In pursuit of a more powerful test for ulcer risk assessment, it has been shown that pre-ulceration areas become warm and inflamed around 35 days before the ulcer actually appears [5,8,14]. This warning sign is invisible to the naked eye, but visible with thermal imaging (Figure 1) and could give doctors enough lead time to intervene and prevent the ulcer from forming. Our team member Kayla moved to India on a Fulbright Fellowship and collected thermal images of patients at all different stages of ulcer development. She then trained a deep learning model to classify high risk patients (those having previous history of ulceration) from low-risk ulcer patients (those without diabetic neuropathy) based on thermal scans alone, with the model performing with 83% accuracy on the balanced dataset.

**Figure 1:**
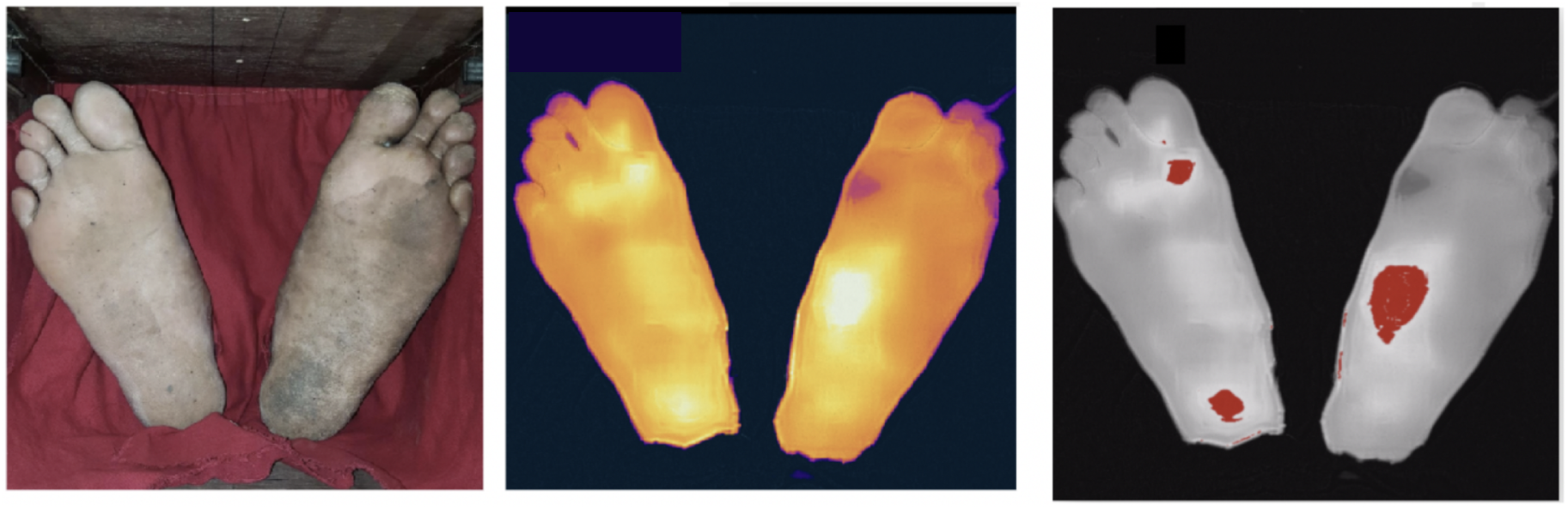
Comparison of visible spectrum image (left), FLIR thermal camera scan (center), and highlights of warmest areas of the feet. These hot spots may be indicative of a developing wound.

### 1.1 Overall problem plan

#### Improve upon 83% baseline accuracy

- Facilitate learning in our models by using segmentation or detection of the feet to then be used as input to our ulcer risk classification model. We hypothesize that this will help the model learn faster by only being presented with the regions of interest (the pixels pertaining to the feet).
- We plan to do this by experimenting with other pre-trained models and approaches (ResNet50, Transformer models, DFTNet and increased compute power).
- Understand if there is benefit to augmenting the input with a visible spectrum image. We will experiment with 1) early fusion: concatenating the images as input and 2) late-stage fusion: concatenating outputs of parallel models in the last layer of the model.

#### Increase the model’s explainability

- Our idea is to understand if the thermal images can also give accurate prediction of the patient’s neuropathy score (current standard of care for diagnosing neuropathy). This would make the model’s risk prediction clinically interpretable if it’s accompanied by a predicted neuropathy score.
  A. Predict a patient’s monofilament score: use of a cat-like whisker, tapping 10 designated areas on the foot for detecting loss of sensation on the patient’s foot. We’ve labeled the patients neuropathy status as positive if a patient cannot feel at least 6/10 spots.
  B. Predict a patient’s biothesiometer score: instrument used to identify vibration sensory loss, another component of peripheral sensory neuropathy. The instrument is operated from 0-50V of vibration. We’ve labeled the patient’s neuropathy status as positive if a patient cannot feel prior to 25V of vibration.

### 1.2 A summary of our results

By 1) experimenting with different models 2) implementing detection of feet in images as preprocessing to the model, and 3) augmenting input to the model with visual images, we were able to achieve over 99.4% accuracy on our test set in predicting a patient’s risk for ulcer development. In addition, our attempts to improve explainability of the model by predicting a patient’s neuropathy risk (current standard of care) achieved 83.9% accuracy in predicting monofilament score and 90.9% accuracy in predicting biothesiometer scores.

## 2 Related Work

### Thermal Prediction classification

Traditional methods like Support Vector Machines (SVM), Adaboost, Random Forest, k-nearest neighbors (k-NN) etc. are very common classifiers for image classification. In [20], Ribeiro has applied SVM to the redundancy reduced diabetes data and achieved a high accuracy of 98.47%. In recent years, deep learning based methods have shown more promising performance in image classification. In [21], six deep CNN models have been used in thermograms image classification including MobilenetV2[22], Resnet18[23], Resnet50[23], DenseNet201[24], InceptionV3[25] and VGG19[26]. Among these models, MobilenetV2 outperforms others for a two feet thermogram image classification. In [7], Cruz-Vega et al. proposed the DFTNet for the five-level classification of diabetic foot thermograms, although their results were not tied to a clinical classification of foot ulcer risk.

More recently, Transformers have garnered more attention in the image classification field. In [27], Vision Transformer based on self-attention has been applied in several image recognition benchmarks (ImageNet, CIFAR-100, VTAB, etc.) and attains great results even compared with the state-of-the-art CNNs. Swin Transformer based on ViT [8] is a hierarchical Transformer with shifted windows when computing representation and has achieved excellent performance on several image classification, object detection, and semantic segmentation tasks.

The idea to use deep learning begin to classify thermograms of diabetic feet was first published by Goyal et al [19] who built a CNN architecture which they call DFUNet (Diabetic Foot Ulcer Net) built to classify thermal scans of patients with diabetes into one of 5 classes. Namely, they separate control patient thermal scans from 4 categories of thermal variation. These thermal categories simply bin from a histogram stratifying the variation seen in a patient’s thermal scan (0 = least variance in temperature, 5 = highest variance). However, they give no discussion of the clinical relevance of these 5 classes (healthy vs diabetic vs neuropathic vs ulceration). DFUNet performance was evaluated by the model’s accuracy at classifying each pair of classes from each other (1 vs 5, 1 vs 4, 1 vs 3 etc). Because our model’s output will identify the clinical risk category of the patient, it will therefore have more clinical relevance be poised to suggest if a patient should receive intervention.

### Detection

Object detection locates and categorizes objects of interest in an image - challenging and core problem in the field of computer vision. Nowadays, object detection in deep learning is mainly divided into two categories, i.e., two-stage and one-stage object detection algorithms. Two-stage detectors are proposal-based detectors. It will first generate region proposals and then do classification on these proposals. In R-CNN[30], the regional proposals are produced based on Selective Search. Based on it, Fast R-CNN[31] then adopted the ROI pooling layer to convert feature maps with different sizes to a fixed size and enables end-to-end detector training on shared convolutional features which also shows compelling accuracy and speed[32]. In Faster R-CNN, the Region Proposal Network is developed to generate proposal regions. As for one-stage detectors, they are all region-free methods, category probabilities and position coordinates of objects are generated in one stage. SSD[33] uses multiscale feature maps, by combining these feature maps with different resolutions, it could adapt to do detections on targets in different sizes. YOLOv3[34] adopts residual blocks and also uses multiscale feature maps to get the bounding box output. As for YOLOv4[35], it has a high accuracy in real-time target detection and meanwhile finds the balance between accuracy and speed. More recently, anchor-free objection detectors have developed as well. FCOS[36] is an anchor box free and proposal-free one-stage object detector. It uses the Center-ness to reduce the number of low-quality bounding boxes which are far away from targets.

## 3 Data

Our team member Kayla spent a year living in India as a Fulbright scholar studying how healthcare workers identify and care for patients at high-risk for ulceration. She worked to identify and quantify the early thermal warning sign on patients’ feet in hopes that it could improve time to intervention for patients with diabetes. She captured thermal scans from March through May of 2019. The Ethics committee/IRB of CMC Vellore Hospital gave ethical approval for this work (IRB Min. No. 11644; dated 28.11.2018).

### Images (thermal and visible)

The dataset include 241 patients’ who either were consented for study through the endocrinology deparment of CMC Vellore (Tamil Nadu, India) or the outpatient clinic at CMC Vellore - Chittoor campus (Andhra Pradesh, India). The study population includes 5 different clinical categories:

1. No diabetes - low ulcer risk
2. Diabetes, no neuropathy - low ulcer risk
3. Diabetes and neuropathy - unknown ulcer risk
4. Diabetes, ulcerated patients - high risk for new ulceration
5. Diabetes, neuropathy patients with previously healed ulcer - high risk for re-ulceration

For each patient, we recorded their status of diabetes, neuropathy score (monofilament 0-10, biothesiometer 0-50V), and whether there was currently and ulcer present. In addition, we captured a phone camera picture of the plantar region of their feet as well as a thermal scan using an FLIPOne Pro thermal camera [44]. These images were 640 × 480 pixels (an example of low and high-risk patients shown in **Figure 2**.)

**Figure 2:**
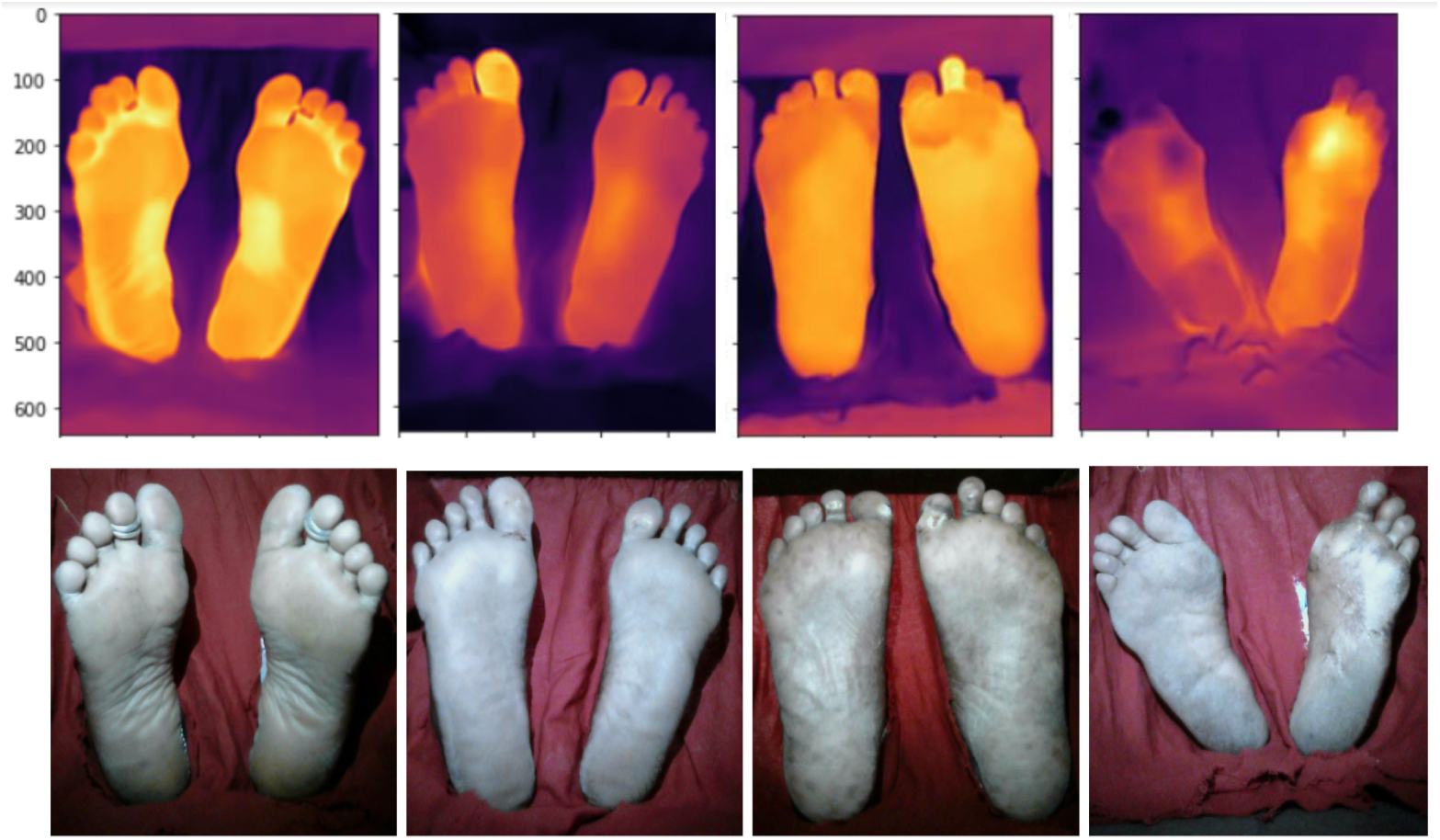
Left most patient does not have neuropathy (i.e. at low risk for ulceration). The symmetry of their thermal profile is contrasted against the right three patients who all have neuropathy but no visible indication of an ulcer.

The dataset includes 112 scans of low-risk feet, 68 images of neuropathy feet, and 55 scans of ulcerated feet. For risk classification, groups 1 and 2 are labeled low risk whereas groups 4 and 5 are labeled high risk. Group 3 is eliminated from this classification task because their risk is unknown.

### Clinical data

Relevant to our experiments here, we also have access to the patients’ neuropathy test scores: both their monofilament and biothesiometer readings. Monofilament scores are on a scale from 0-10, with 6 or fewer being high risk for ulceration. Bioethesiometers scores are on a scale from 0-50, with greater than 25 being high risk for ulceration. Therefore, we’ve assigned 2 additional labels alluding to the monofilament and biothesiometer scores.

## 4 Methods

The goal of our project is to predict diabetic foot ulceration risk based on the thermal and visible spectrum images. To achieve the goal, we aim to provide a method consisting of ROI (region of interest) extraction and classification. The high-level process flow of our end-to-end model is shown in Fig 3.

**Figure 3:**
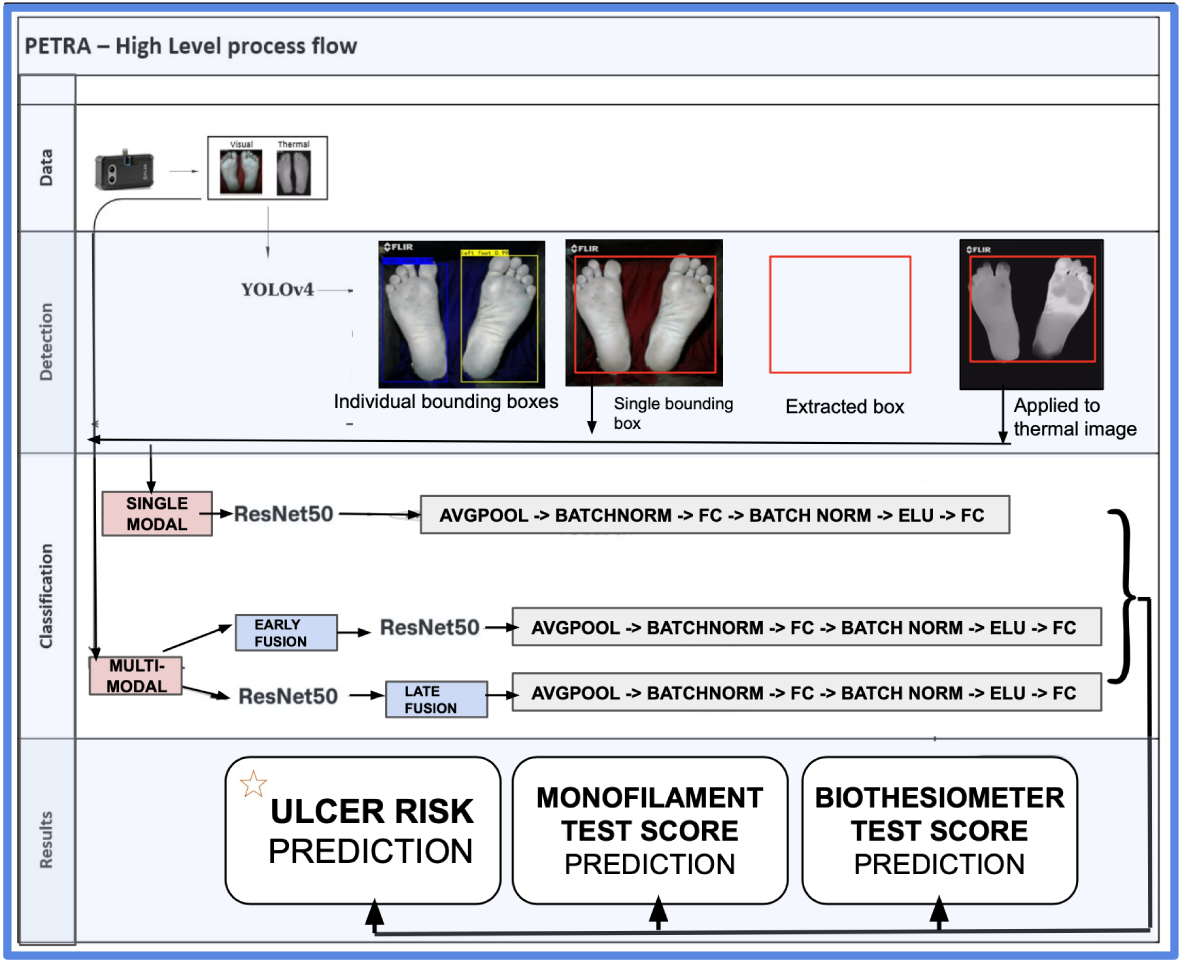
High level process diagram beginning with image capture (thermal and visible) using the FLIR thermal camera. These images and their foot annotations were used to train a YOLOv4 network to detect feet in the images. These identified boxes were used as input to train our ResNet50 algorithm on 3 classification tasks: monofilament risk score, biothesiometer risk score and overall ulcer risk.

### 4.1 Detection

Since the background in thermal images may have no contribution to the classification for the risk of having a new ulcer, we plan to extract a bounding box of each foot in hopes that inputting only the ROI of the feet will yield better performance. After thoroughly reviewing the data, we found that some images have limited contrast between the feet and the background, compromising performance of an unsupervised segmentation algorithm. For example, in some visible spectrum images, there are shadows on the feet and in thermal images, the temperature in some parts of the feet are similar to the background. Therefore, instead of creating a segmentation algorithm, we plan to train a YOLOv4[35] detection network to extract bounding boxes of the feet in our images. We will then experiment with using these bounding boxes as input to our classification models.

### 4.2 Classification

For the classification, we proposed a Convolution Neural Network based on ResNet-50[40] which is pre-trained on ImageNet. The input is first fed to the backbone to get the feature map. Then we use the Average Pooling layer to reduce the size of the extracted feature map from 2048 × 7 × 7 to 2048 × 1. After this, a two-cascade fully connected layer is adopted for classification. In this two-cascade fully connected layer, we also use Batch Normalization and ELU[41] to enhance the robustness of our proposed model. The structure of this proposed model is shown in Fig 4A.

**Figure 4:**
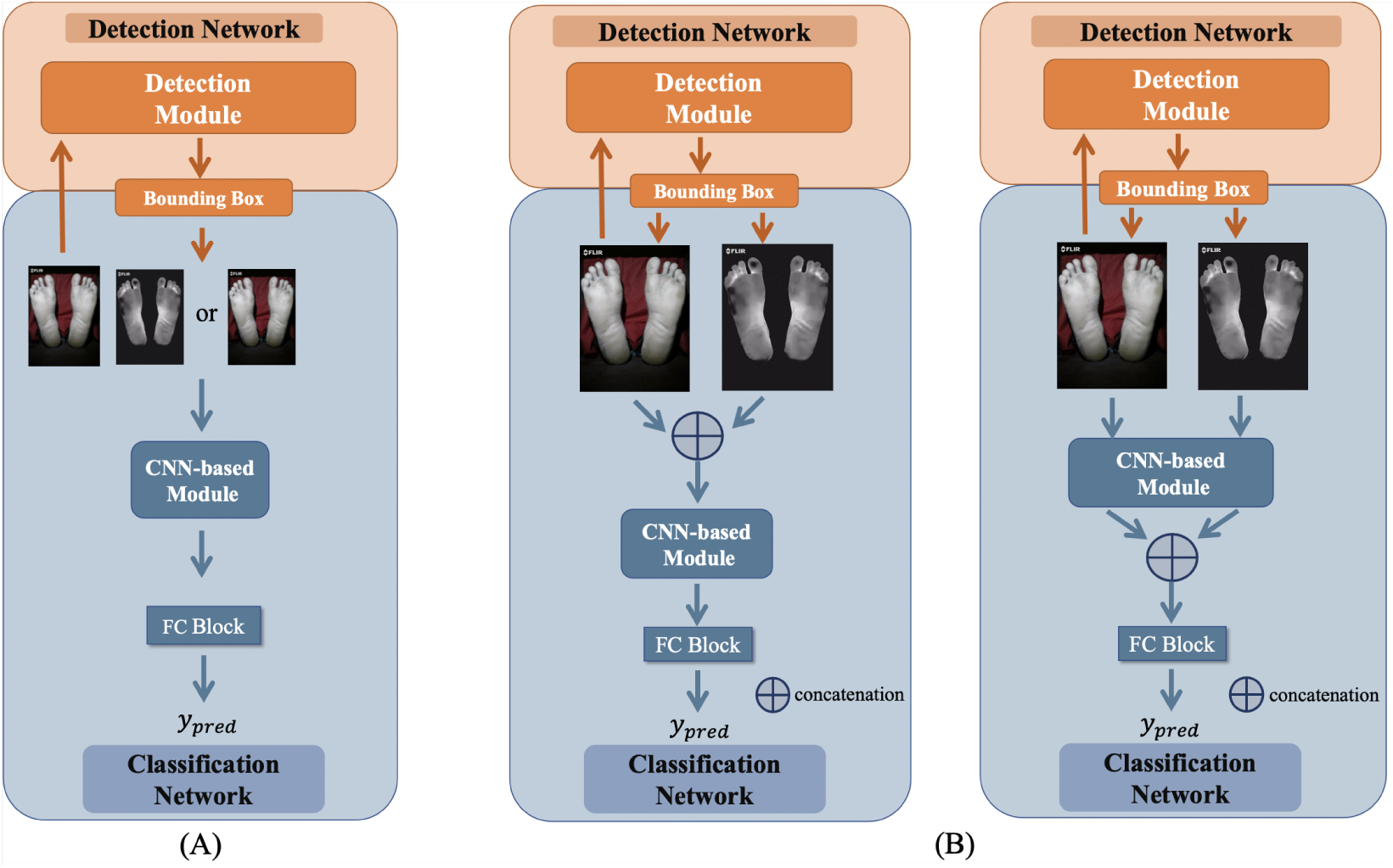
Structure of A) single modality model and B) early fusion and late fusion multi-modality models

Since we have both thermal images and visual spectrum images for each patient, we also developed multi-modal models in our experiments, implemented using two different strategies: early fusion and late fusion. For early fusion, the thermal images and visual spectrum images are concatenated at the beginning which doubles the size of the input channel. For the late fusion, the two images are first fed into the shared-weight backbone separately, and the outputs are then concatenated along the dimension of channels. These concatenated features then become input of the Average Pooling layer, followed by the two-cascade fully connected layers to get the final prediction. These two different multi-modal models are shown in Fig 4B.

We also used the TRIPOD checklist as suggested by the EQUATOR network when writing our report and is attached as supplemental material [43].

## 5 Results

### 5.1 Detection

In total, we have 212 visual spectrum images (train: 169, val: 43) which we used to train the YOLOv4 model in order to detect the left and right feet. During training, we used Adam optimizer and pretrained YOLO v4 model. First, we froze the backbone and trained the free layers for 20 epochs (with an initial lr = 0.001, decay: λ = 0.94). Then we unfreezed the backbone parameters and trained the whole network for 100 epochs (lr = 0.0001, decay: λ = 0.94). All the inputs are resized to 416 × 416. The output bounding boxes of YOLO v4 are shown in Figure 5. The detection network achieves very high mAP and also good IoU performance shown in Figure 5 and Figure 6.

**Figure 5:**
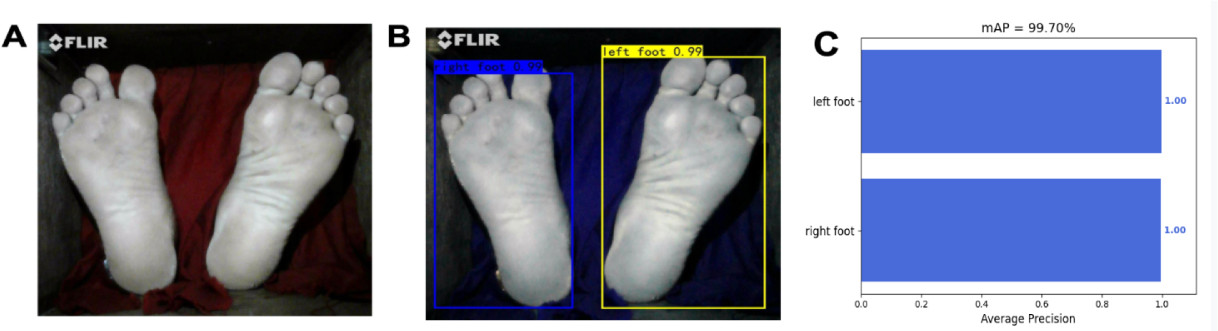
A) Raw Visual Spectrum images B) YOLOv4 predictions on foot presence after training C) AP score of left and right foot

**Figure 6:**
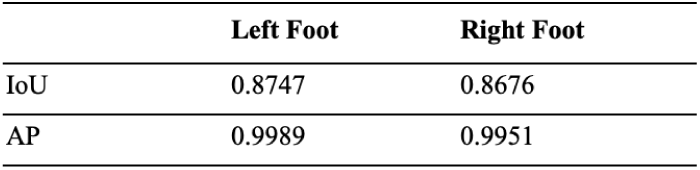
IoU and AP for left and right foot in the validation set from the trained YOLOv4

### 5.2 Classification

#### Selection of backbone for classification

To begin with, we have chosen three candidate models as backbone which are DFTNet, ResNet50 and Swin Transform. Among them, we will select the best performing model as our backbone.

We ground our approach beginning with DFTNet which was developed to identify visible warning signs of ulceration. Although this classification is obvious and not particularly helpful since the ulcer is already visible, this is a good baseline because the pre-ulceration areas or “hot-spots” that show up in thermal images as shown in Figure 1 look geometrically very similar to ulcers that have broken through the skin. In addition, they are located in the same area as the already-formed ulcers. Therefore, beginning with the DFTNet architecture is a good baseline. As for ResNet50, it not only has an accessible pretrained model to use, but it also shows very promising results in other classification tasks. As for transformers, they have now become a hot trend in image classification tasks and Swin Transformer also shows good performances in classification and has accessible pre-trained models which could also be used in our task.

To pick the best model, we evaluate it’s baseline performance on predicting monofilament scores using visual spectrum images. Since Resnet50 outperformed the other two models, we chose it as our backbone, results displayed in Figure 7. We believe such poor performance of the Swin Transformer may be due to the small size of the dataset.

**Figure 7:**
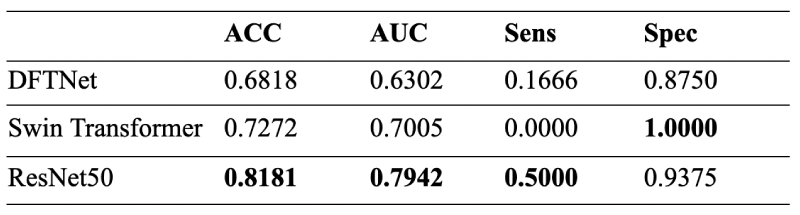
Comparison of results from three candidate models for monofilament prediction task based on original VS images

#### Prediction tasks

In addition to our main task predicting ulcer risk based on the thermal images, to enhance the interpretability of this model, we also conducted experiments on other tasks to predict Monofilament and Biothesiometer scores which are related to patient ulcer risk and used to evaluate the neuropathy of a patient. By showing the risk predicted label combined with monofilament and biothesiometer predicted labels, we believe it could help the clinicians better understand the risk prediction results of our model. In all three training tasks, we adopted pretrained ResNet50, trained by an Adam optimizer (initial lr = 0.0001, decay: *λ* = 150*/*(150 + *epoch*)). We also resize all the input to 256 × 256 and adopted focal loss [42] to balance positive and negative samples.

#### Risk Prediction

Our classes consist of low risk for ulceration (Group 1, 2) and high-risk patients (Group 4, 5). Namely, we leave out Group 3 (neuropathy group) since their risk for ulceration is an ambiguous group and not well understood. Once we train the model to differentiate between the well-defined low-risk and high-risk groups, we can then use this model to infer the risk score of the neuropathy patients (Group 3). The hypothesis is that those neuropathy patients which get classified as high-risk (i.e. their thermal scans are very similar to patients already with an ulcer) would be flagged as high-risk. Those patients who are classified to the low-risk category therefore have thermal scans are similar to those without neuropathy and without diabetes - two groups known to be at low-risk for ulceration.

For the risk prediction task, we had 167 patients from Groups 1, 2, 4, 5 in our datasets. The split of the dataset with labels is shown in Figure 8.

**Figure 8:**
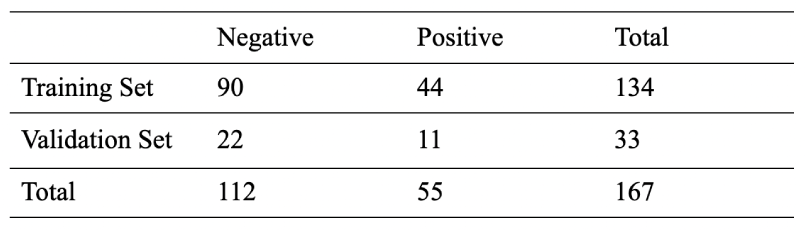
splits of the datasets for risk prediction task

In order to understand what value the visual spectrum images may have on our classification task, we’ve design an ablation study to distill the ability of either image, or early or late combination of the two, in prediction of ulcer risk. For single modality (SM), we conducted experiments on thermal (T), as well as visual spectrum (VS) images separately. As for multi-modality (MM), we carried out both early fusion and late fusion MM models. In each experiment, we used 5-Fold cross-validation. The results of the average are shown in Figure 9.

**Figure 9:**
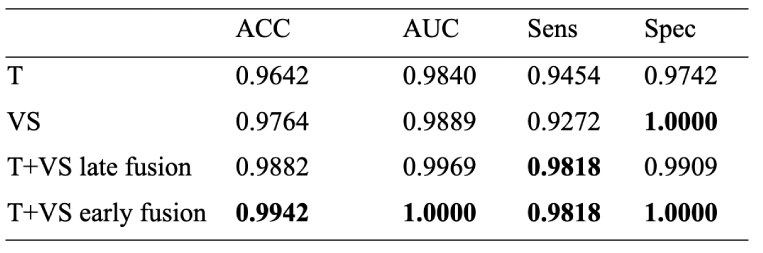
5-Fold Cross-Validation average results of the risk predictions (T: Thermal images VS: Visual spectrum images)

From these results, both thermal and visual spectrum show good performance. Combining the two did improve the results of the SM model, with early fusion performing better than the late fusion model.

#### Monofilament

For the monofilament prediction task, we used the same architecture chosen for ulcer risk classification. In this task, 200 patients in total had monofilament scores recorded. The split of the dataset with labels is shown in Figure 10.

**Figure 10:**
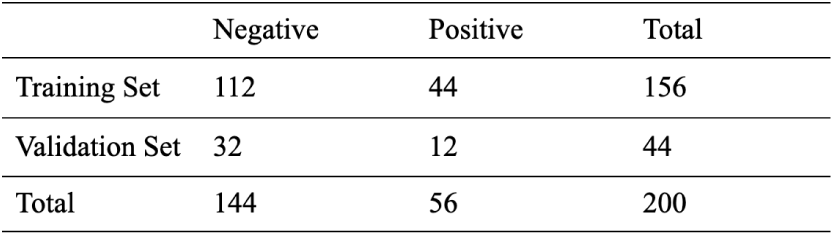
splits of the datasets for monofilament prediction task

For the ablation study, we have conducted several experiments for the SM and MM models to distill whether applying bounding boxes of feet to the input(s) improve performance. The results are shown in Figure 10.

From Figure 11, we can see that applying bounding boxes improved performance, confirming our hypothesis that the background doesn’t help with learning. In addition, visual spectrum images outperformed thermal images. However both early and late fusion MM models didn’t improve results.

**Figure 11:**
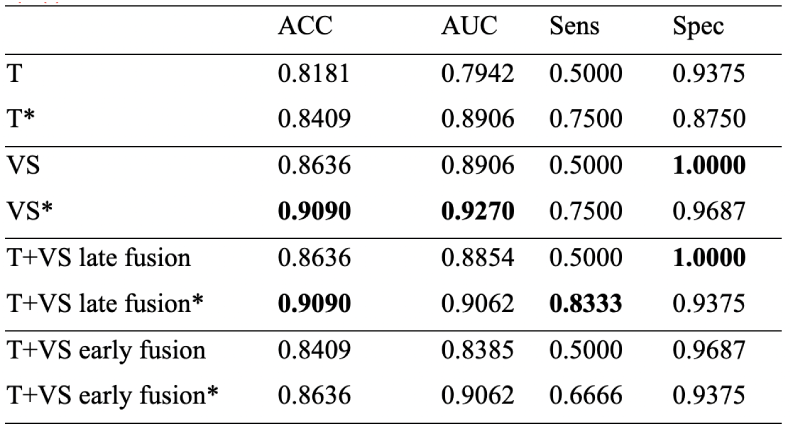
ablation study results of the monofilament prediction task The symbol * means applying bounding boxes which are acquired from the trained YOLO v4 to the input(s)

#### Biothesiometer

We used the same architecture chosen for ulcer risk classification. In this task, included the 139 patients that had a recorded biothesiometer reading. The split of the dataset with labels is shown in Figure 12. Following the same pattern of the monofilament ablation experiments, we carried out SM and MM model experiments. We also compared whether applying bounding boxes would improve performance. From the results (Figure 13), we see that thermal images with bounding boxes showed best performance but that both MM models didn’t improve the performance.

**Figure 12:**
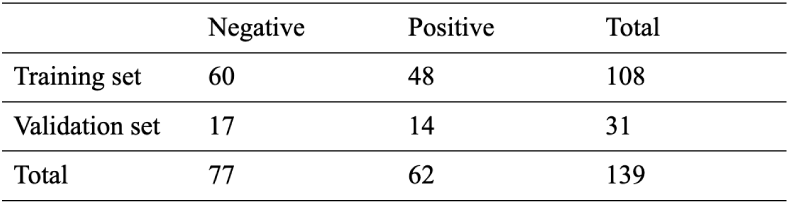
splits of the datasets for Biothesiometer prediction task

**Figure 13:**
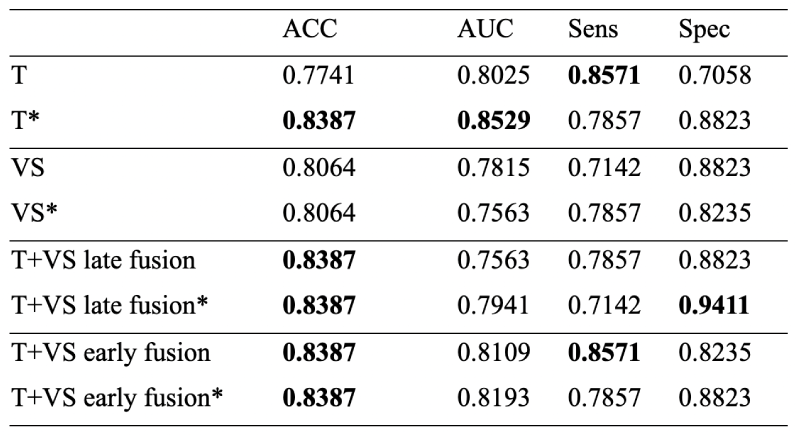
ablation study results of the biothesiometer prediction task, the symbol * means applying bounding boxes which are acquired from the trained YOLO v4 to the input(s)

## 6 Conclusion

For various experiments we performed, our best accuracy is 99.43% when classifying high-risk patients from low-risk patients, resulting from the early fusion MM model. Using this model, we were able to perform inference on what the neuropathy patient’s risk scores would be (Figure 14). Interestingly, a few patients did in-fact score high, while a majority were classified as low-risk. The limitation here is that since we are unable to follow-up with these patients, we do not know their outcome to validate our prediction. For our future work, we would like to tconduct a longitudinal study where patients are tracked over time, collecting time series data which will allow the prediction task to be a time-until-ulceration prediction.

**Figure 14:**
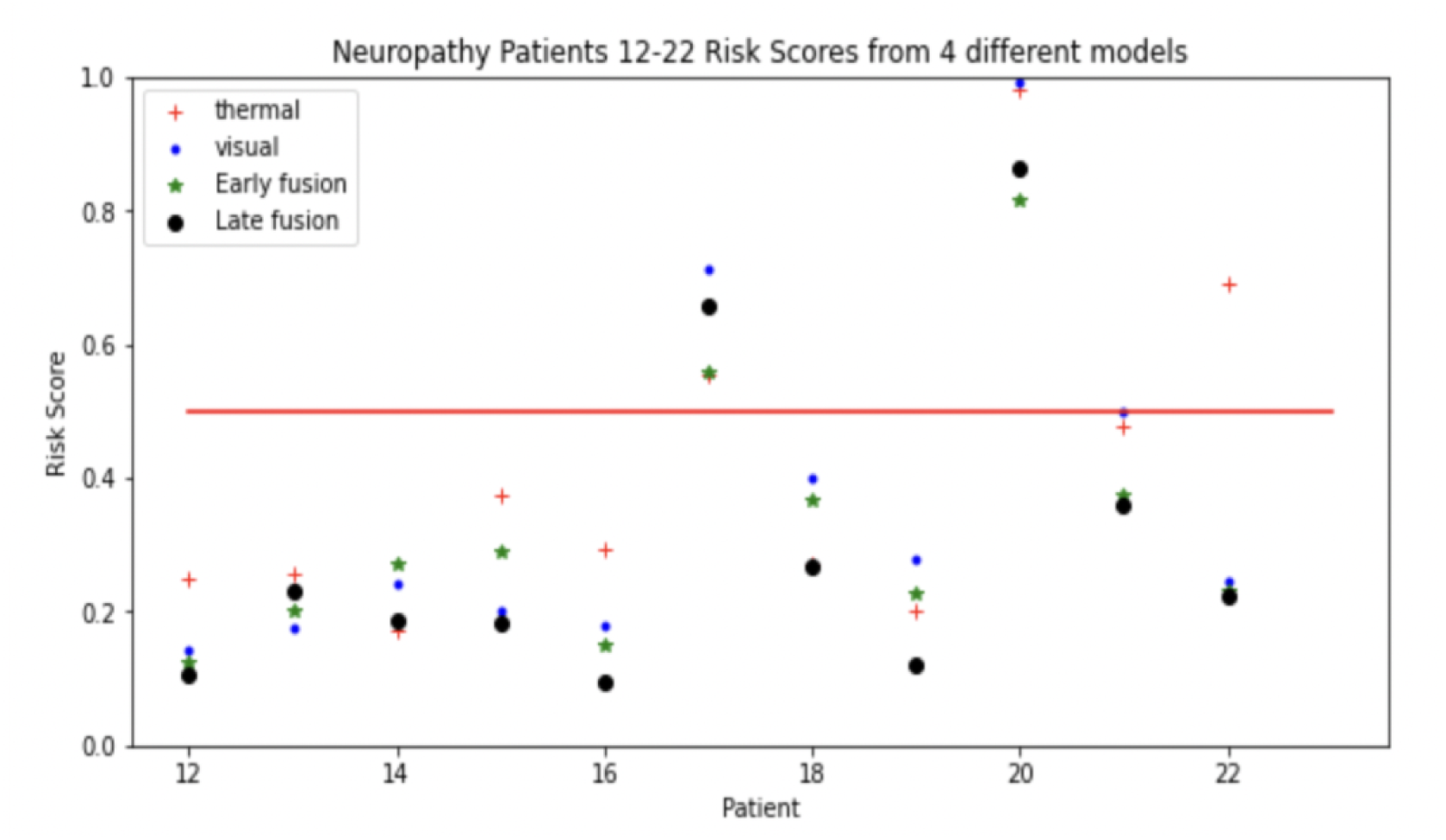
Ulcer risk predictions made on subset of neuropathy patients for whom their ulcer is ambiguous. Note the agreeance between models and the clear deviation of patients 17 and 20 from the rest of the patients.

Despite our attempt to increase interpretability of our model by predicting monofilament and biothesiometer test scores to give transparency regarding the patient’s predicted neuropathy score, both early and late fusion multi-modality models didn’t show better performance as the risk classification task. We think it might be because of our chosen fusion mechanism. Since early or late fusion are just simple concatenation operations, we might need better algorithms to take thorough advantage of both modalities. For example we could add attention mechanisms in our future study.

One other major learning from this project is that visible spectrum images performed just as well if not better than the thermal images on all our classification tasks. If we could validate this in a larger study to show that our model is robust and generalizable, this would be revolutionary. This risk stratification would therefore not be limited by who have access to a thermal camera - theoretically, a patient could snap a picture from home to receive more understanding of their risk which could prompt them to seek care while the ulcer can still be prevented.To this end, we believe this technology has the potential to enable preventative care for patients with diabetes and ultimately prevent millions of amputations around the world.

## Supporting information

Completed Equator Network, TRIPOD checklist

## Data Availability

All data produced in the present study may be made available upon reasonable request to the authors.

## Author Declarations

The study was approved by the CMC Vellore IRB ethics committee. IRB Min. No. 11644; dated 28.11.2018

## References

[1] “Diabetes facts and figures.” International Diabetes Federation https://www.idf.org/aboutdiabetes/what-isdiabetes/facts-figures.html

[2] Lin, X., Xu, Y., Pan, X. et al. Global, regional, and national burden and trend of diabetes in 195 countries and territories: an analysis from 1990 to 2025. Sci Rep 10, 14790 (2020). https://doi.org/10.1038/s41598-020-71908-9

[3] Reyzelman AM, Koelewyn K, Murphy M, Shen X, Yu E, Pillai R, Fu J, Scholten HJ, Ma R. Continuous Temperature-Monitoring Socks for Home Use in Patients With Diabetes: Observational Study. J Med Internet Res. 2018 Dec 17;20(12):e12460. doi: 10.2196/12460. PMID: 30559091; PMCID: PMC6315272.

[4] Singh N, Armstrong DG, Lipsky BA. Preventing foot ulcers in patients with diabetes. JAMA 2005; 293: 217–228.

[5] Armstrong DG, Holtz-Neiderer K, Wendel C, et al. Skin temperature monitoring reduces the risk for diabetic foot ulceration in high-risk patients. Am J Med 2007;120:1042–6.doi:10.1016/j.amjmed.2007.06.028

[6] Lavery LA, Higgins KR, Lanctot DR, Constantinides GP, Zamorano RG, Athanasiou KA, Armstrong DG, Agrawal CM. Preventing diabetic foot ulcer recurrence in high-risk patients: use of temperature monitoring as a self-assessment tool. Diabetes Care. 2007 Jan;30(1):14–20. doi: 10.2337/dc06-1600. PMID: 17192326.

[7] Cruz-Vega, I.; Hernandez-Contreras, D.; Peregrina-Barreto, H.; Rangel-Magdaleno, J.d.J.; Ramirez-Cortes, J.M. Deep Learning Classification for Diabetic Foot Thermograms. Sensors 2020, 20, 1762.

[8] Liu, Z., et al. Swin Transformer: Hierarchical Vision Transformer using Shifted Windows. (2021).

[9] Rasmussen, B. S., Froekjaer, J., Joergensen, L. B., Halekoh, U. Yderstraede, K. B. Validation of a new imaging device for telemedical ulcer monitoring. Skin Res. Technol. 21, 485–492 (2015).

[10] Foltynski, P., Ladyzynski, P. Wojcicki, J. M. A new smartphone-based method for wound area measurement. Artif. Organs 38, 346–352 (2014).

[11] https://www.nature.com/articles/s41598-017-09828-4

[12] https://en.wikipedia.org/wiki/Watershed (image processing) Watershed By Flooding

[13] Barnes, R., Lehman, C., Mulla, D., 2014. Priority-flood: An optimal depression-filling and watershed labeling algorithm for digital elevation models. Computers Geosciences 62, 117–127

[14] Zhou Z, Siddiquee M, Tajbakhsh N, et al. UNet++: Redesigning Skip Connections to Exploit Multiscale Features in Image Segmentation[J]. IEEE Transactions on Medical Imaging, 2020, 39(6):1856–1867.

[15] Liu, C.; van der Heijden, F.; Klein, M.E.; van Baal, J.G.; Bus, S.A.; van Netten, J.J. Infrared dermal thermography on diabetic feet soles to predict ulcerations: a case study. In Proceedings of the SPIEBiOS, Advanced Biomedical and Clinical Diagnostic Systems XI, San Francisco, CA, USA, 2–7 February 2013.

[16] Etehadtavakol, M.; Ng, E.; Kaabouch, N. Automatic segmentation of thermal images of diabetic-at-risk feet using the snakes algorithm. Infrared Phys. Technol. 2017, 86, 66–76.

[17] Barnes, R., Lehman, C., Mulla, D., 2014. Priority-flood: An optimal depression-filling and watershed labeling algorithm for digital elevation models. Computers Geosciences 62, 117–127

[18] Zhang, K.; Zhang, L.; Song, H.; Zhou W. Active contours with selective local or global segmentation: A new formulation and level set method. Image Vision Comput. 2010, 28, 668–676.

[19] Goyal, M., et al. “DFUNet: Convolutional Neural Networks for Diabetic Foot Ulcer Classification.” (2017).

[20] Ribeiro, Á.C.; Barros, A.K.; Santana, E.; Príncipe, J.C. Diabetes classification using a redundancy reduction preprocessor. Research on Biomed. Eng. 2015, 31, 97–106

[21] Khandakar A, Chowdhury MEH, Ibne Reaz MB, et al. A machine learning model for early detection of diabetic foot using thermogram images. Comput Biol Med. 2021;137:104838. doi:10.1016/j.compbiomed.2021.104838

[22] Sandler, M., Howard, A., Zhu, M., Zhmoginov, A., Chen, L. C.. (2018). MobileNetV2: Inverted Residuals and Linear Bottlenecks. 2018 IEEE/CVF Conference on Computer Vision and Pattern Recognition (CVPR). IEEE.

[23] He, K., Zhang, X., Ren, S., Sun, J.. (2016). Deep residual learning for image recognition. IEEE. 8

[24] K. Adam, I. I. Mohamed and Y. Ibrahim, “A Selective Mitigation Technique of Soft Errors for DNN Models Used in Healthcare Applications: DenseNet201 Case Study,” in IEEE Access, vol. 9, pp. 65803–65823, 2021, doi: 10.1109/ACCESS.2021.3076716.

[25] Szegedy, C., Vanhoucke, V., Ioffe, S., Shlens, J., Wojna, Z.. (2016). Rethinking the Inception Architecture for Computer Vision. 2016 IEEE Conference on Computer Vision and Pattern Recognition (CVPR) (pp.2818–2826). IEEE.

[26] Simonyan, K., Zisserman, A.. (2014). Very deep convolutional networks for large-scale image recognition. Computer Science.

[27] Dosovitskiy, A., Beyer, L., Kolesnikov, A., Weissenborn, D., Houlsby, N.. (2020). An image is worth 16×16 words: transformers for image recognition at scale.

[28] Manu Goyal, Neil D Reeves, Satyan Rajbhandari, and Moi Hoon Yap. Robust methods for real-time diabetic foot ulcer detection and localization on mobile devices. IEEE Journal of Biomedical and Health Informatics, 23(4):1730–1741, July 2019.

[30] Ross Girshick, Jeff Donahue, Trevor Darrell, and Jitendra Malik. Rich feature hierarchies for accurate object detection and semantic segmentation. In Proceedings of the IEEE Conference on Computer Vision and Pattern Recognition (CVPR), pages 580–587, 2014. 2, 4

[31] RossGirshick. FastR-CNN. In Proceeding soft he IEEE International Conference on Computer Vision (ICCV), pages 1440–1448, 2015.

[32] Shaoqing Ren, Kaiming He, Ross Girshick, and Jian Sun. Faster R-CNN: Towards real-time object detection with region proposal networks. In Advances in Neural Information Processing Systems (NIPS), pages 91–99, 2015. 2

[33] Wei Liu, Dragomir Anguelov, Dumitru Erhan, Christian Szegedy, Scott Reed, Cheng-Yang Fu, and Alexander C Berg. SSD: Single shot multibox detector. In Proceedings of the European Conference on Computer Vision (ECCV), pages 21–37, 2016. 2, 11

[34] Joseph Redmonand Ali Farhadi. YOLOv3:Anincremental improvement. arXiv preprint 1804.02767, 2018. 2, 4, 7, 11

[35] Bochkovskiy, A., C. Y. Wang, and H. Liao. “YOLOv4: Optimal Speed and Accuracy of Object Detection.” (2020).

[36] Zhi Tian, Chunhua Shen, Hao Chen, and Tong He. FCOS: Fully convolutional one-stage object detection. In Proceedings of the IEEE International Conference on Computer Vision (ICCV), pages 9627–9636, 2019. 2

[37] Chien-Yao Wang, Hong-Yuan Mark Liao, Yueh-Hua Wu, Ping-Yang Chen, Jun-Wei Hsieh, and I-Hau Yeh. CSPNet: A new backbone that can enhance learning capability of cnn. Proceedings of the IEEE Conference on Computer Vision and Pattern Recognition Workshop (CVPR Workshop), 2020. 2, 7

[38] Kaiming He, Xiangyu Zhang, Shaoqing Ren, and Jian Sun. Spatial pyramid pooling in deep convolutional networks for visual recognition. IEEE Transactions on Pattern Analysis and Machine Intelligence (TPAMI), 37(9):1904–1916, 2015. 2, 4, 7

[39] Shu Liu, Lu Qi, Haifang Qin, Jianping Shi, and Jiaya Jia. Path aggregation network for instance segmentation. In Proceedings of the IEEE Conference on Computer Vision and Pattern Recognition (CVPR), pages 8759–8768, 2018. 1,2,7

[40] Kaiming He, Xiangyu Zhang, Shaoqing Ren, and Jian Sun. Deep residual learning for image recognition. In Proceedings of the IEEE International Conference on Computer Vision (ICCV), pages 9627–9636, 2019. 2

[41] Clevert, Djork-Arné, T. Unterthiner, and S. Hochreiter. “Fast and Accurate Deep Network Learning by Exponential Linear Units (ELUs).” Computer Science (2015).

[42] Lin, T. Y., et al. “Focal Loss for Dense Object Detection.” IEEE Transactions on Pattern Analysis Machine Intelligence PP.99(2017):2999–3007.

[43] Collins GS, Reitsma JB, Altman DG, Moons KG. Transparent reporting of a multivariable prediction model for individual prognosis or diagnosis (TRIPOD): The TRIPOD statement.

[44] FLIROne Pro Thermal camera: https://www.flir.com/products/flir-one-pro

