## Supplementary material for "Preventing diabetic foot ulcers in low resource settings using Pedal Elevated Temperature Risk Assessment (PETRA)": Completed Equator Network, TRIPOD checklist

### Reporting checklist for prediction model development/validation.

Based on the TRIPOD guidelines.

#### Instructions to authors

Complete this checklist by entering the page numbers from your manuscript where readers will find each of the items listed below.

Your article may not currently address all the items on the checklist. Please modify your text to include the missing information. If you are certain that an item does not apply, please write "n/a" and provide a short explanation.

Upload your completed checklist as an extra file when you submit to a journal.

In your methods section, say that you used the TRIPODreporting guidelines, and cite them as:

Collins GS, Reitsma JB, Altman DG, Moons KG. Transparent reporting of a multivariable prediction model for individual prognosis or diagnosis (TRIPOD): The TRIPOD statement.

|  |  | Reporting Item | Page Number |
| --- | --- | --- | --- |
| **Title** |  |  |  |
|  | [#1](https://www.goodreports.org/reporting-checklists/tripod/info/#1) | Identify the study as developing and / or validating a multivariable prediction model, the target population, and the outcome to be predicted. | 2-3 |
| **Abstract** |  |  |  |
|  | [#2](https://www.goodreports.org/reporting-checklists/tripod/info/#2) | Provide a summary of objectives, study design, setting, participants, sample size, predictors, outcome, statistical analysis, results, and conclusions. | 1 |
| **Introduction** |  |  |  |
|  | [#3a](https://www.goodreports.org/reporting-checklists/tripod/info/#3a) | Explain the medical context (including whether diagnostic or prognostic) and rationale for developing or validating the multivariable prediction model, including references to existing models. | 2-5 |
|  | [#3b](https://www.goodreports.org/reporting-checklists/tripod/info/#3b) | Specify the objectives, including whether the study describes the development or validation of the model or both. | 5 |
| **Methods** |  |  |  |
| Source of data | [#4a](https://www.goodreports.org/reporting-checklists/tripod/info/#4a) | Describe the study design or source of data (e.g., randomized trial, cohort, or registry data), separately for the development and validation data sets, if applicable. | 4 |
| Source of data | [#4b](https://www.goodreports.org/reporting-checklists/tripod/info/#4b) | Specify the key study dates, including start of accrual; end of accrual; and, if applicable, end of follow-up. | 4 |
| Participants | [#5a](https://www.goodreports.org/reporting-checklists/tripod/info/#5a) | Specify key elements of the study setting (e.g., primary care, secondary care, general population) including number and location of centres. | 4 |
| Participants | [#5b](https://www.goodreports.org/reporting-checklists/tripod/info/#5b) | Describe eligibility criteria for participants. | 4 |
| Participants | [#5c](https://www.goodreports.org/reporting-checklists/tripod/info/#5c) | Give details of treatments received, if relevant | n/a |
| Outcome | [#6a](https://www.goodreports.org/reporting-checklists/tripod/info/#6a) | Clearly define the outcome that is predicted by the prediction model, including how and when assessed. | 7-8 |
| Outcome | [#6b](https://www.goodreports.org/reporting-checklists/tripod/info/#6b) | Report any actions to blind assessment of the outcome to be predicted. | n/a |
| Predictors | [#7a](https://www.goodreports.org/reporting-checklists/tripod/info/#7a) | Clearly define all predictors used in developing or validating the multivariable prediction model, including how and when they were measured | 7-9 |
| Predictors | [#7b](https://www.goodreports.org/reporting-checklists/tripod/info/#7b) | Report any actions to blind assessment of predictors for the outcome and other predictors. | n/a |
| Sample size | [#8](https://www.goodreports.org/reporting-checklists/tripod/info/#8) | Explain how the study size was arrived at. | n/a |
| Missing data | [#9](https://www.goodreports.org/reporting-checklists/tripod/info/#9) | Describe how missing data were handled (e.g., complete-case analysis, single imputation, multiple imputation) with details of any imputation method. | n/a |
| Statistical analysis methods | [#10a](https://www.goodreports.org/reporting-checklists/tripod/info/#10a) | If you are developing a prediction model describe how predictors were handled in the analyses. | n/a |
| Statistical analysis methods | [#10b](https://www.goodreports.org/reporting-checklists/tripod/info/#10b) | If you are developing a prediction model, specify type of model, all model-building procedures (including any predictor selection), and method for internal validation. | 7-9 |
| Statistical analysis methods | [#10c](https://www.goodreports.org/reporting-checklists/tripod/info/#10c) | If you are validating a prediction model, describe how the predictions were calculated. | 7-9 |
| Statistical analysis methods | [#10d](https://www.goodreports.org/reporting-checklists/tripod/info/#10d) | Specify all measures used to assess model performance and, if relevant, to compare multiple models. | 7-9 |
| Statistical analysis methods | [#10e](https://www.goodreports.org/reporting-checklists/tripod/info/#10e) | If you are validating a prediction model, describe any model updating (e.g., recalibration) arising from the validation, if done | n/a |
| Risk groups | [#11](https://www.goodreports.org/reporting-checklists/tripod/info/#11) | Provide details on how risk groups were created, if done. | n/a |
| Development vs. validation | [#12](https://www.goodreports.org/reporting-checklists/tripod/info/#12) | For validation, identify any differences from the development data in setting, eligibility criteria, outcome, and predictors. | n/a |
| **Results** |  |  |  |
| Participants | [#13a](https://www.goodreports.org/reporting-checklists/tripod/info/#13a) | Describe the flow of participants through the study, including the number of participants with and without the outcome and, if applicable, a summary of the follow-up time. A diagram may be helpful. | 8 |
| Participants | [#13b](https://www.goodreports.org/reporting-checklists/tripod/info/#13b) | Describe the characteristics of the participants (basic demographics, clinical features, available predictors), including the number of participants with missing data for predictors and outcome. | 8 |
| Participants | [#13c](https://www.goodreports.org/reporting-checklists/tripod/info/#13c) | For validation, show a comparison with the development data of the distribution of important variables (demographics, predictors and outcome). | n/a |
| Model development | [#14a](https://www.goodreports.org/reporting-checklists/tripod/info/#14a) | If developing a model, specify the number of participants and outcome events in each analysis. | 8 |
| Model development | [#14b](https://www.goodreports.org/reporting-checklists/tripod/info/#14b) | If developing a model, report the unadjusted association, if calculated between each candidate predictor and outcome. | n/a |
| Model specification | [#15a](https://www.goodreports.org/reporting-checklists/tripod/info/#15a) | If developing a model, present the full prediction model to allow predictions for individuals (i.e., all regression coefficients, and model intercept or baseline survival at a given time point). | n/a |
| Model specification | [#15b](https://www.goodreports.org/reporting-checklists/tripod/info/#15b) | If developing a prediction model, explain how to the use it. | n/a |
| Model performance | [#16](https://www.goodreports.org/reporting-checklists/tripod/info/#16) | Report performance measures (with CIs) for the prediction model. | n/a |
| Model-updating | [#17](https://www.goodreports.org/reporting-checklists/tripod/info/#17) | If validating a model, report the results from any model updating, if done (i.e., model specification, model performance). | n/a |
| **Discussion** |  |  |  |
| Limitations | [#18](https://www.goodreports.org/reporting-checklists/tripod/info/#18) | Discuss any limitations of the study (such as nonrepresentative sample, few events per predictor, missing data). | n/a |
| Interpretation | [#19a](https://www.goodreports.org/reporting-checklists/tripod/info/#19a) | For validation, discuss the results with reference to performance in the development data, and any other validation data | n/a |
| Interpretation | [#19b](https://www.goodreports.org/reporting-checklists/tripod/info/#19b) | Give an overall interpretation of the results, considering objectives, limitations, results from similar studies, and other relevant evidence. | 8-10 |
| Implications | [#20](https://www.goodreports.org/reporting-checklists/tripod/info/#20) | Discuss the potential clinical use of the model and implications for future research | 9-10 |
| **Other information** |  |  |  |
| Supplementary information | [#21](https://www.goodreports.org/reporting-checklists/tripod/info/#21) | Provide information about the availability of supplementary resources, such as study protocol, Web calculator, and data sets. | n/a |
| Funding | [#22](https://www.goodreports.org/reporting-checklists/tripod/info/#22) | Give the source of funding and the role of the funders for the present study. | 10 |

The TRIPOD checklist is distributed under the terms of the Creative Commons Attribution License CC-BY. This checklist was completed on 26. June 2022 using <https://www.goodreports.org/>, a tool made by the [EQUATOR Network](https://www.equator-network.org) in collaboration with [Penelope.ai](https://www.penelope.ai)
